# Ambulance or corridor? The association between site-level use of ambulance ‘ramping’ as Emergency Department escalation areas and 28-day mortality in admitted patients: a secondary analysis of the UNCORKED study

**DOI:** 10.64898/2026.07.25.26358912

**Authors:** Ryan D Mchenry, Fraser Birse, Benjamin Clarke, Tom Roberts, the Trainee Emergency Research Network

**Author notes:** **Corresponding Author**, Ryan D McHENRY ScotSTAR, Scottish Ambulance Service, Paisley, UK, School of Health and Wellbeing, University of Glasgow, Glasgow, UK. **Funding** The study was funded by the Royal College of Emergency Medicine. Grant number RCEM24_SG_4.

## Abstract

**Background and importance:** Emergency Department (ED) crowding is an increasing public health concern, with evidence of harms to patients, the public and healthcare systems. When the number of patients requiring emergency care exceeds the capacity of EDs, operational decisions must be made regarding the best place of care for patients. It is not known if there is an association between place of escalation area care (for example ‘ramping’ in an ambulance, or in an ED corridor) and patient outcomes.

**Objective(s):** This study aimed to assess the association between the relative proportion of all escalation area care that a site provided in an ambulance (the Ambulance:Escalation Index) and all-cause 28-day mortality.

**Design:** A secondary analysis of a prospective cohort study. Adult patients ≥16 years attending EDs in England, Wales and Northern Ireland in March 2025.

**Intervention or exposure (if any):** The Ambulance:Escalation Index, a site-level indicator of the proportion of all time in escalation area care provided in an ambulance.

**Outcome measures and analysis:** The profile of site-level ambulance use was presented descriptively. Multivariable survival analysis was used to assess the primary outcome, all-cause 28-day mortality.

**Main results:** Of 131 EDs using escalation area, 82 (62.6%) used ambulances as a place of escalation area care. There was a significant association between a site’s increasing use of ambulances as escalation areas and mortality; for each 5% increase in the proportion of a site’s total escalation area care delivered in ambulances, there was a 2.1% increase in the hazard of death by 28 days (HR 1.021, 95% CI 1.002-1.041, p=0.032).

**Conclusion:** Emergency Department crowding is associated with poor outcomes irrespective of where departments are forced to deliver care; however this study suggests that there is excess mortality where escalation area care is more often delivered in ambulances.

## Introduction

Emergency Department (ED) crowding is an increasing international public health concern, [1,2] with evidence of harm to patients, [3] and clinicians. [4] The causes of crowding are well-established, [5] and in contemporary emergency care, the major reason for crowding are output issues, those of ‘exit block’ or poor access to inpatient beds for those requiring admission. [1,6]

When the number of patients requiring emergency care exceed the capacity of EDs, in conjunction with regional ambulance services, departments are forced to choose how they will care for patients. This has led to the use of ‘escalation areas’, such as corridors, doubled-up cubicles, or ‘ramping’ in ambulances, where patients wait outside of EDs for longer than necessary for handover to in-hospital teams. [7,8] The UNCORKED study reported the incidence and outcomes of ED-based corridor care in the UK in March 2025. [6] Since then, the UK government committed to ending corridor care, [9] and simultaneously has limited the time for which ambulances may wait before transferring patients to the care of EDs to 45 minutes, with an aim of less than 15 minutes. [10]

Long waits in ambulances outside EDs are widely accepted to harm both the patient held in the ambulance and patients in the community awaiting an ambulance response, [7] and long ED waits and escalation area care are known to increase length of stay and mortality. [3,11] However, a recent systematic review considering the individual harms of ambulance ‘ramping’ has concluded that evidence of harm is limited,[12] and there is no evidence comparing these two undesirable models of care against each other: when escalation care cannot be avoided, it is unknown whether holding patients in ambulances or within the ED is associated with worse outcomes.

At the time of the UNCORKED study, whether patients waited in ambulances or in ED escalation areas was at the discretion of local decision makers. This variation in local policy provides a natural experiment: sites facing comparable escalation pressure resolved it in systematically different ways, allowing the two models of care to be compared directly. Specifically, it allows assessment of:

1. The extent to which hospitals preferentially cared for patients in ambulances rather than in-hospital ED escalation areas, and
2. Whether mortality differed between patients admitted from sites adopting these different models of care.

This secondary analysis of the UNCORKED dataset aimed to report the number of UK EDs using an ambulance-based model of escalation care, as opposed to an in-hospital ED-based model, or a relative proportion of each, and to estimate the association between an operationally-significant decision, the proportion of a site’s escalation area time spent in ambulances, and 28-day mortality among patients admitted from the ED.

## Methods

The methodology for the study has been reported in the primary analyses [6] and was conducted as a prospective multicentre observational study led by the Royal College of Emergency Medicine’s (RCEM) Trainee Emergency Research Network (TERN). This analysis included patients recruited from 134 type 1 EDs (providing consultant-led, 24-hour services with full resuscitation facilities) in England, Wales and Northern Ireland. Although part of the study reported data from 13 EDs in Scotland, due to differing national approaches to regulatory approval of recruitment via waived consent, Scotland did not collect patient level data.

Eligible patients were recruited at five predetermined snapshots over 14 days, chosen to give a representative spread of ED activity based on published attendance data: [13]

1. 12:00, 03/03/2025 (Monday)
2. 07:00, 06/03/2025 (Thursday)
3. 16:00, 08/03/2025 (Saturday)
4. 19:00, 10/03/2025 (Monday)
5. 23:59, 12/03/2025 (Wednesday)

### Participants

Trained clinical research nurses and Emergency Medicine clinicians prospectively identified all patients present in the ED during the snapshots.

Demographics were recorded for all patients admitted from the ED, and these patients were subsequently screened at 28 days to determine their mortality outcome. Patients were recruited via waived consent. This study reports the data for adult patients ≥16 years and excludes paediatric-only Emergency Departments.

There is no universally agreed definition for an escalation area. The following definition was provided to sites: ‘any area not routinely used unless the capacity of the usual ED geographical footprint is exceeded.’ Sites were asked to assign each escalation area to one of the following categories:

- Ambulance queueing to offload for >15 minutes.
- Repurposed clinical area.
- Non-clinical area (e.g. hospital corridor).
- Doubled-up cubicle.

Patients in the waiting room were considered to be in a non-clinical escalation area if there was objective evidence that they would be in a standard ED cubicle if one were available (e.g. receiving intravenous medication, and/or awaiting an inpatient bed). Other than queuing ambulances and pre-hospital cohort areas, only patients in escalation areas under the care of the ED team were included. The central study team liaised with sites to identify relevant areas within their EDs.

The mean total escalation area time for each ED was calculated in hours, as the sum of all patients’ time in any escalation area for that department, divided by the number of patients in each ED across all snapshots. The mean *ambulance* escalation area time for each department was calculated in the same way, restricted to only those waits in an ambulance delayed >15 minutes for handover. An Ambulance:Escalation Index was then constructed, by dividing the total ambulance escalation area time by the total escalation area time for each department, giving a ratio describing the relative use of ambulance escalation areas for each department, regardless of the total burden of escalation area care.

### Outcomes

The proportion of sites using ambulance-based escalation care, and the extent of that use is reported descriptively. The primary outcome was 28-day all-cause mortality among patients admitted from the ED, with the site-level Ambulance:Escalation Index as the exposure.

### Data collection

Local study teams prospectively identified all patients present in the ED during snapshots. They subsequently used electronic health records (EHR), department management systems and in-department observation to determine patients’ disposition from the ED. Patient demographics were collected from patient records. At 28-days, hospital and GP records were examined for evidence of patient death. Data were entered using REDCap electronic data capture tools. [14]

### Statistical methods and analysis

The characteristics of the included EDs, and patients recruited, were described as counts, or median and interquartile range (IQR).

Spearman’s rank correlation was used to assess whether a site’s relative use of ambulance escalation areas was associated with its total burden of escalation area care (mean total escalation area time), to establish that the index measures the composition of escalation care rather than its volume.

The association between the Ambulance:Escalation Index and 28-day all-cause mortality was estimated with a Cox proportional hazards model. The exposure was parameterised in two parts: a binary indicator of any use of ambulance escalation areas by the site, and a continuous dose term, the Ambulance:Escalation Index. The model adjusted for patient age (natural cubic spline with four degrees of freedom), sex, hospital trauma-receiving designation (Local Emergency Hospital, Trauma Unit or Major Trauma Centre), recruitment snapshot, site admission rate, and the site’s total burden of escalation area care (mean total escalation area time per recruited patient). Because the index and the total burden share components, this adjustment means the Ambulance:Escalation Index is independent of total escalation burden: the association with shifting escalation area time towards ambulances, holding the total escalation burden per patient constant.

The baseline hazard was stratified by diagnosis group. As the exposure varies only between sites, inference was based on robust standard errors clustered at the hospital level. Hazard ratios (HRs) for the index are presented per 5 percentage-point absolute increase for interpretability. A directed acyclic graph demonstrating the assumed causal structure is available as Supplementary Figure 1.

The proportional hazards assumption was assessed using scaled Schoenfeld residuals, and the functional form of continuous covariates using martingale residuals. [15] The linearity of the dose-response association was assessed with a penalised-spline Cox model (restricted maximum likelihood smoothing) including a hospital-level random effect, judged by the effective degrees of freedom of the exposure smooth. The stability of the dose-response estimate was assessed by leave-one-out analysis by site. [16] The data were analysed on a complete-case basis and statistical significance was set at p<0.05. Hospitals reporting no escalation area use were excluded. Analyses were conducted in R (version 4.5.0) using the survival and mgcv packages. [17] The study is reported according to STROBE guidance. [18]

### Patient and Public Involvement

Patients and the public were involved in an RCEM-facilitated event where support was expressed for the utility, design and dissemination of this research.

### Registry & Ethnical Approval

The study was prospectively registered at the ISCTRN (ref: ISRCTN16396025). The study received ethical approval from the Brighton and Sussex Research Ethics Committee (24/LO/0837) and registered at ISCTRN (ISRCTN16396025).

## Results

The study included 134 EDs, of which 131 (97.8%) reported the use of any escalation area at any time and so were included for analysis. These sites had a median total mean per-patient escalation area time of 1.76 hours (IQR 0.67-3.53 hours). 37.4% (n=49) of sites reported no use of ambulance escalation areas. The median Ambulance:Escalation Index of those sites that used ambulance escalation spaces was 0.05 (IQR 0.01-0.12) (Figure 1).

**Figure 1.**
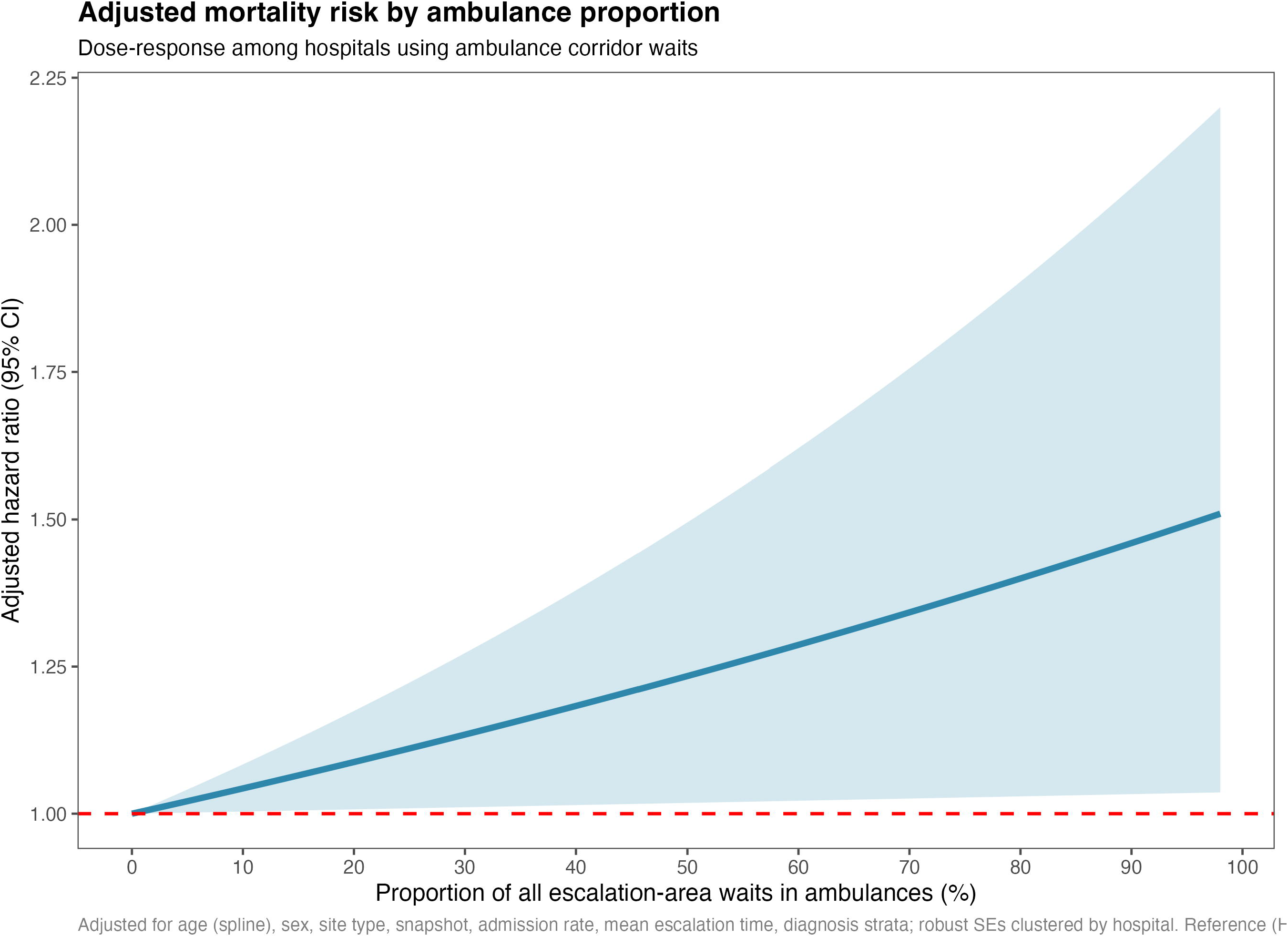
Characteristics of hospital sites, by the mean escalation time per patient, the proportion of that escalation time in ambulance. The admission rates for sites and the total number of patients recruited (volume) are additionally indicated by colour and size respectively.

18,744 patients, admitted from the Emergency Department, were included. The median age was 72 (IQR 55-83), and 9,036 (48%) were Male. 1,240 patients (6.6%) had died at 28-day follow-up. Mortality was highest in those departments with highest Ambulance:Escalation Index, with 8.9% mortality for ED admissions in those departments using ambulances for >15% of their total escalation area time, compared to 6.4% for those without ambulance escalation area use.

The characteristics of the EDs and admitted patients, stratified by the Ambulance:Escalation Index, are presented in Table 1.

**Table 1.**
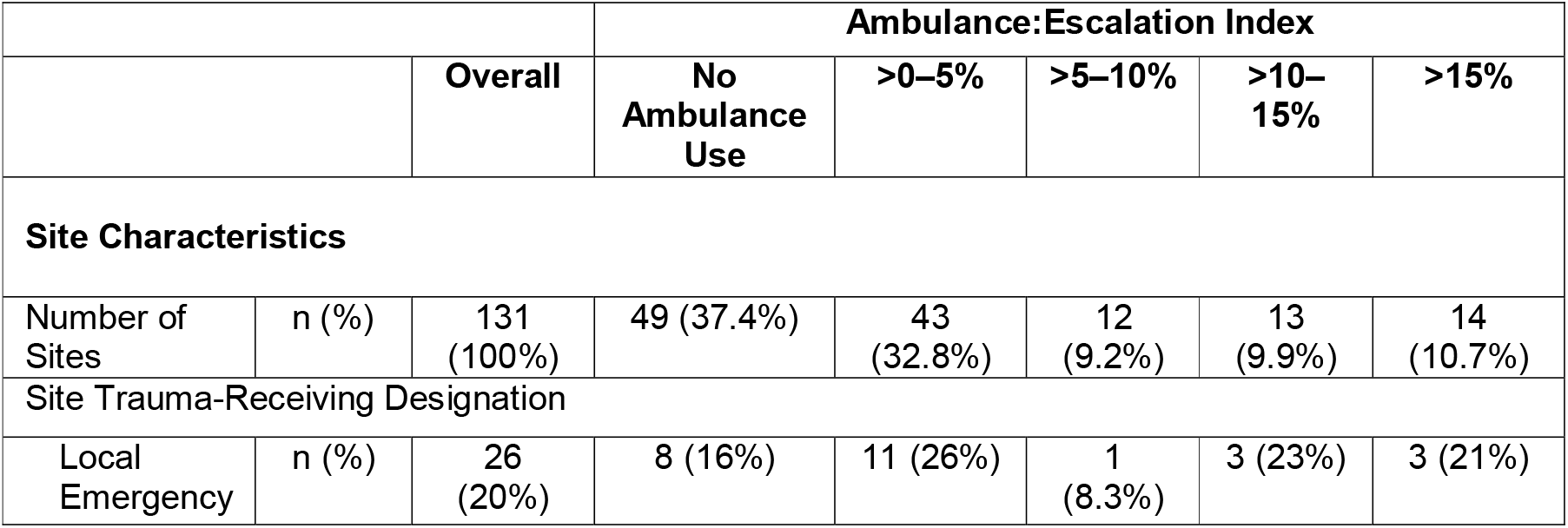
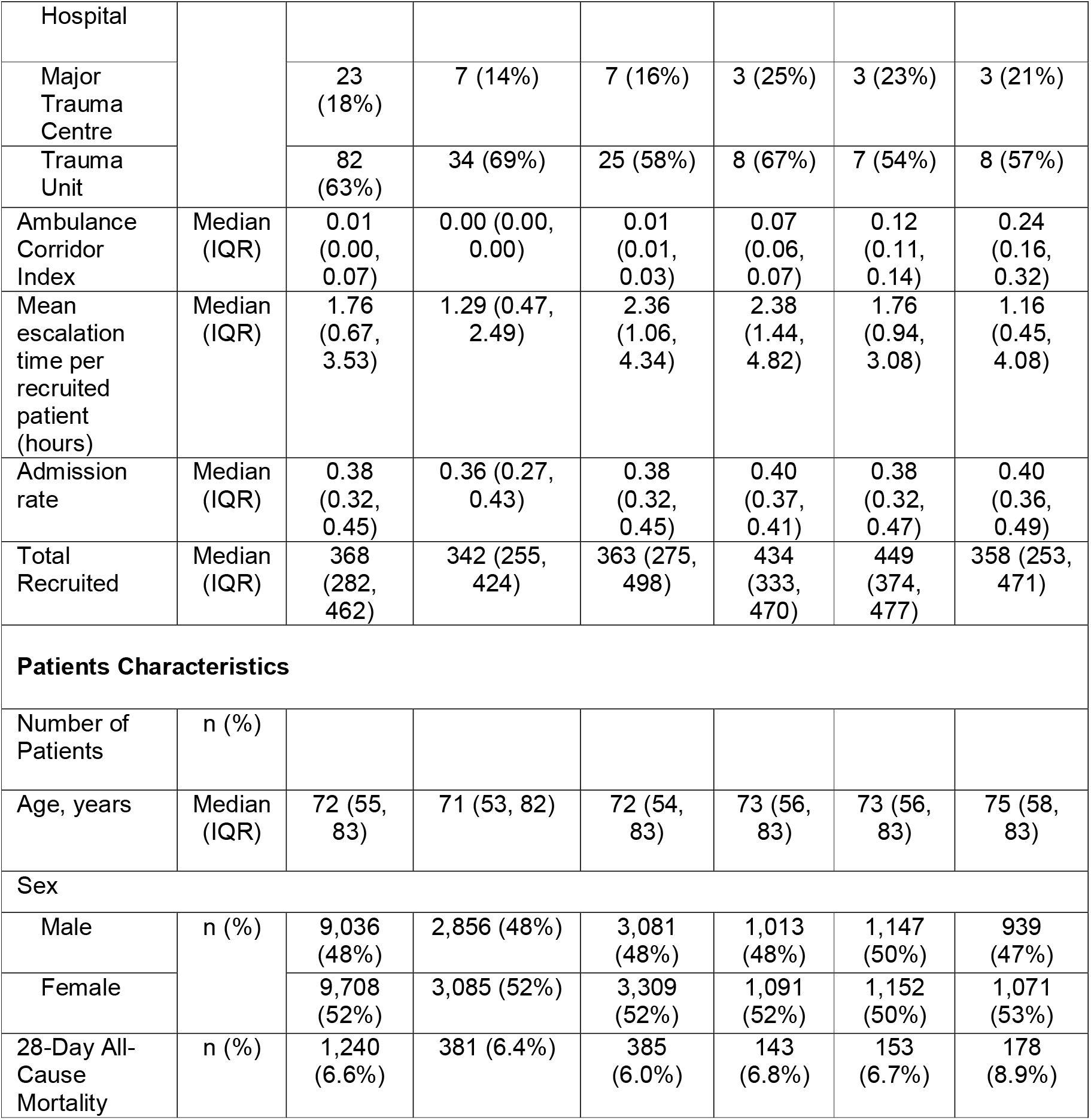
Site and patient characteristics by site Ambulance:Escalation Index (proportion of each site’s total escalation-area time spent in ambulances)

For those sites using ambulance escalation, there was no significant correlation between a site’s relative use of ambulance escalation areas and its mean total escalation area time (Spearman’s rho = -0.149, p=0.183), supporting the index as a measure of the composition of escalation care rather than its overall burden.

There was no association between the binary use, or not, of ambulance escalation areas and mortality (HR 0.957, 95% CI 0.808-1.134, p=0.612). Among sites using ambulance escalation areas, increasing relative use was associated with increased mortality: each 5 percentage-point absolute increase in the Ambulance:Escalation Index was associated with a 2.1% relative increase in the hazard of death (HR 1.021, 95% CI 1.002-1.041, p=0.032). The model demonstrated visual proportionality. The linearity of this relationship was confirmed with visual inspection of the penalised-spline Cox model and was robust to leave-one-out analysis (Supplementary Figures 2 & 3). The full output of the modelling is presented in Table 2, and the association between the Ambulance:Escalation Index and mortality is shown in Figure 2.

**Table 2.** Results from the univariable and multivariable Cox proportional hazards survival analysis assessing the effect of the Ambulance:Escalation Index on all-cause 28-day mortality.

|  | Univariable |  | Multivariable |  |
| --- | --- | --- | --- | --- |
|  | HR (95%CI) | p value | HR (95% CI) | p value |
| Site use of ambulance escalation areas | 1.009 (0.833–1.222) | 0.928 | 0.957 (0.808–1.134) | 0.612 |
| Ambulance:Escalation Index (for 5% increase, for sites using ambulance escalation areas) | 1.020 (0.998–1.043) | 0.077 | 1.021 (1.002–1.041) | 0.032 |
| Mean escalation time (for each additional hour) | 1.034 (1.015–1.053) | <0.001 | 1.025 (1.005–1.044) | 0.012 |
| Site admission rate (for 5% increase) | 1.060 (1.025–1.096) | <0.001 | 1.024 (0.992–1.057) | 0.137 |
| Sex |  |  |  |  |
| Male |  |  | Reference | — |
| Female | 0.760 (0.679–0.851) | <0.001 | 0.747 (0.666–0.838) | <0.001 |
| Site Trauma-Receiving Designation |  |  |  |  |
| Local Emergency Hospital |  |  | Reference | — |
| MTC | 1.007 (0.742–1.366) | 0.966 | 1.161 (0.882–1.527) | 0.288 |
| Trauma Unit | 1.163 (0.906–1.493) | 0.235 | 1.212 (0.948–1.550) | 0.125 |
| Age |  |  |  |  |
| 50 vs 25 years (spline contrast) | 8.312 (4.223–16.359) | <0.001 | 7.136 (3.649–13.953) | <0.001 |
| 65 vs 25 years (spline contrast) | 21.617 (10.917–42.803) | <0.001 | 16.338 (8.313–32.110) | <0.001 |
| 80 vs 25 years (spline contrast) | 38.463 (19.653–75.277) | <0.001 | 29.225 (15.063–56.699) | <0.001 |

**Figure 2.**
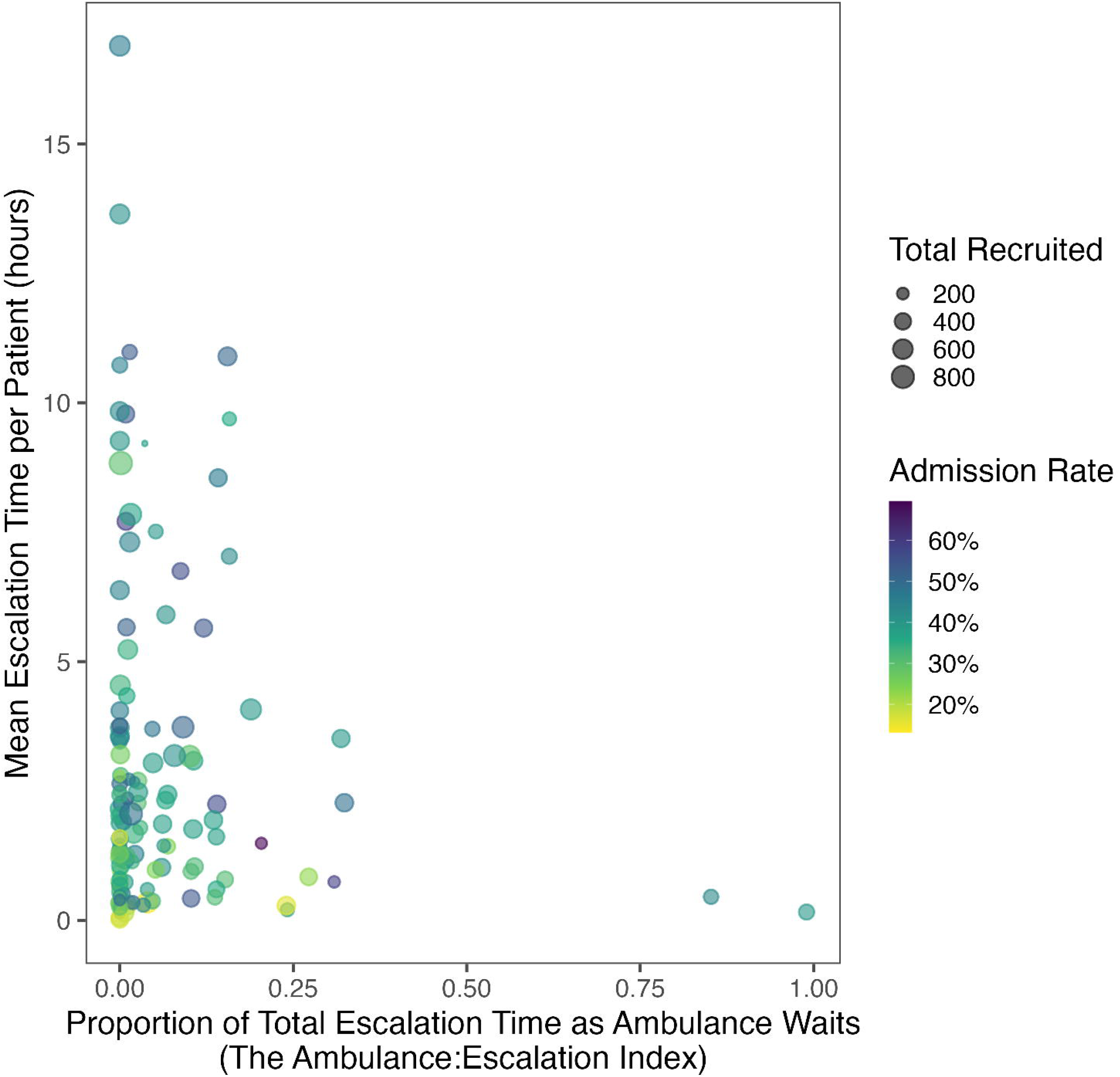
Plot demonstrating the association between the Ambulance:Escalation Index and the adjusted Hazard Ratio. The line is the central estimate and the shaded area demonstrates the 95% confidence intervals.

### HR – Hazard Ratio

Hazards are stratified by diagnostic group and additionally adjusted for recruitment snapshot. For the univariable model, each row group is estimated from a separate Cox model containing only that variable, with robust standard errors clustered by hospital; not stratified by diagnosis group. The model has been constructed to assess the causal effect of the Ambulance:Escalation Index, and the results of other covariates in the model should not be interpreted in the same way.

## Discussion

### Statement of principal findings

In this secondary analysis of 131 UK EDs, the use of ambulances as escalation areas was widespread: three in five sites held patients in ambulances for more than 15 minutes, and at one in five sites ambulances accounted for over 10% of all escalation area time. Among sites that did use ambulance escalation area care, mortality rose steadily with the share of escalation care delivered in ambulances: each 5 percentage-point increase in the Ambulance:Escalation Index was associated with a 2.1% relative increase in the hazard of 28-day mortality among admitted patients. This association was independent of the site’s total burden of escalation care, admission rate, case mix and trauma designation. It reflects the relative burden of where escalation care was delivered, not how much of it there was.

This study addresses important gaps, recently identified in the literature, regarding the relative effect of ambulance ‘ramping’ compared to other strategies to manage ED crowding. [12] Its strengths include national scope, prospective recruitment across prespecified representative snapshots, 28-day follow-up through hospital and GP records, and an analysis designed for a site-level exposure: inference was clustered at hospital level across the sites, and the dose-response estimate was robust to leave-one-site-out analysis. Importantly, this analysis did not aim to assess the effect of extra time for patients waiting in ambulance escalation, which may simply reflect additional pressures at each individual site. Instead, it aimed to assess the extent to which decisions surrounding the proportion of escalation area utilisation assigned to ambulance, rather than in-hospital, escalation areas affected patient outcomes.

## Limitations

This is an observational between-site comparison, and because the exposure is a site-level characteristic, no within-site design is possible. Unmeasured differences between sites, particularly in staffing, where long ambulance waits may reflect delays to first clinical assessment due to challenging staffing conditions or inefficient processes.[19] Equally, other operational factors, such as estate footprint or flow management culture could confound the association. The cohort was restricted to admitted patients, if ambulance pressures alter admission thresholds as has been demonstrated in crowded departments previously, [20] this restriction could introduce selection bias. A patient was only deemed to be in an ambulance-based escalation area if their wait exceeded 15 minutes, with no equivalent threshold applied to in-hospital escalation time. While this is pragmatic given the usual requirements for ambulances to spend some time registering their patients with ED teams, it will tend to understate ambulance use at sites with frequent shorter holds. Finally, this remains a hypothesis-generating secondary analysis of a study designed for a different primary question, and causal claims are not supported.

### Strengths and weaknesses in relation to other studies, discussing important differences in results

A previous single-centre study assessed the effect of ambulance ramping on in-patient mortality and length of stay, [7] reporting no significant mortality effect, but was limited by its single-centre design and importantly it assessed individual experience, not the system-wide operational decisions considered in this work. The present analysis supports other evidence of harms from ED crowding.[3,11] It suggests that, beyond the amount of escalation care a patient receives, that operational decisions on the location of care, balancing multiple poor options, carries an additional association with mortality. In this secondary analysis, a causal relationship has been established for neither, but the consistency and dose-dependence of the present association across 131 sites strengthens the case that the choice between ambulance and in-hospital escalation is not neutral and deserves further study.

### Meaning of the study: possible explanations and implications for clinicians, researchers and policymakers

These findings have implications across the international response to ED crowding. For example, current UK guidance requires ambulances to offload within 45 minutes, [10] shifting escalation burden from ambulances into hospitals. This study provides comparative evidence relevant to that choice. Where ambulance escalation care could not be avoided, sites that provided a greater share of their entire escalation area care in ambulances had higher mortality among admitted patients than sites that absorbed it in-hospital. The mechanisms are plausible, as patients held in ambulances have reduced access to ED nursing observation, first assessment, senior review, diagnostics and timely treatment. The harms measured here may be conservative, since they exclude the community harm of ambulances held out of service, which falls entirely on the ambulance model of care.

These results provide early evidence for policies that aim to relieve the burden of ambulance ramping. They are not, however, an endorsement of in-hospital corridor care. The primary results of this study demonstrated such care is itself associated with harm. Likewise, these results do not support any substitute for addressing the flow failures that cause escalation care in the first place.

### Future research

Staggered adoption of ambulance wait limits offers a natural experiment amenable to interrupted time series and difference-in-differences designs, which could test prospectively whether shifting escalation care out of ambulances reduces mortality.

## Conclusion

There is an association between Emergency Departments that provide a greater proportion of their escalation area care in ambulances and all-cause 28-day mortality. Future research should link pre-hospital data to capture the community harms of ambulance holding and identify the mechanisms through which the location of escalation care may influence outcomes. However, none of this should supersede efforts to eliminate escalation area care, which is demonstrably harmful to patients, staff and systems, and where research, policy and resources should be directed.

## Supporting information

STROBE

## Data Availability

All data produced in the present study are available upon reasonable request to the authors and with appropriate ethical and governance consent.

