## Supplementary material for "Ambulance or corridor? The association between site-level use of ambulance ‘ramping’ as Emergency Department escalation areas and 28-day mortality in admitted patients: a secondary analysis of the UNCORKED study": STROBE

STROBE Statement—checklist of items that should be included in reports of observational studies

| **Item No.** | **Recommendation** | **Section** | **Relevant text from manuscript** |
| --- | --- | --- | --- |
| **Title and abstract** |  |  |  |
| 1(a) | Indicate the study’s design with a commonly used term in the title or the abstract | Title & Abstract | “Ambulance or corridor? The association between site-level use of ambulance ‘ramping’ as Emergency Department escalation areas and 28-day mortality in admitted patients: a secondary analysis of the UNCORKED study”  “Design: A secondary analysis of a prospective cohort study. Adult patients ≥16 years attending EDs in England, Wales and Northern Ireland in March 2025.” |
| 1(b) | Provide in the abstract an informative and balanced summary of what was done and what was found | Abstract | “This study aimed to assess the association between the relative proportion of all escalation area care that a site provided in an ambulance (the Ambulance:Escalation Index) and all-cause 28-day mortality.”  “Of 131 EDs using escalation area, 82 (62.6%) used ambulances as a place of escalation area care… for each 5% increase in the proportion of a site’s total escalation area care delivered in ambulances, there was a 2.1% increase in the hazard of death by 28 days (HR 1.021, 95% CI 1.002-1.041, p=0.032).” |
| **Introduction** |  |  |  |
| 2 | Explain the scientific background and rationale for the investigation being reported | Introduction, Paragraphs 1-4 | “Emergency Department (ED) crowding is an increasing international public health concern, with evidence of harm to patients, and clinicians… the major reason for crowding are output issues, those of ‘exit block’…”  “…there is no evidence comparing these two undesirable models of care against each other: when escalation care cannot be avoided, it is unknown whether holding patients in ambulances or within the ED is associated with worse outcomes.” |
| 3 | State specific objectives, including any prespecified hypotheses | Introduction, Paragraphs 5-6 | “This secondary analysis of the UNCORKED dataset aimed to report the number of UK EDs using an ambulance-based model of escalation care, as opposed to an in-hospital ED-based model, or a relative proportion of each, and to estimate the association between an operationally-significant decision, the proportion of a site's escalation area time spent in ambulances, and 28-day mortality among patients admitted from the ED.” |
| **Methods** |  |  |  |
| 4 | Present key elements of study design early in the paper | Methods, Paragraph 1 | “The methodology for the study has been reported in the primary analyses and was conducted as a prospective multicentre observational study led by the Royal College of Emergency Medicine’s (RCEM) Trainee Emergency Research Network (TERN).” |
| 5 | Describe the setting, locations, and relevant dates, including periods of recruitment, exposure, follow-up, and data collection | Methods, Paragraphs 1-2; Results, Paragraph 1 | “This analysis included patients recruited from 134 type 1 EDs (providing consultant-led, 24-hour services with full resuscitation facilities) in England, Wales and Northern Ireland.”  “Eligible patients were recruited at five predetermined snapshots over 14 days… 12:00, 03/03/2025 (Monday) … 23:59, 12/03/2025 (Wednesday)”  “At 28-days, hospital and GP records were examined for evidence of patient death.” |
| 6(a) | Give the eligibility criteria, and the sources and methods of selection of participants | Methods (Participants), Paragraphs 1-3 | “Trained clinical research nurses and Emergency Medicine clinicians prospectively identified all patients present in the ED during the snapshots.”  “Demographics were recorded for all patients admitted from the ED, and these patients were subsequently screened at 28 days to determine their mortality outcome… This study reports the data for adult patients ≥16 years and excludes paediatric-only Emergency Departments.” |
| 6(b) | For matched studies, give matching criteria | N/A | N/A – this is not a matched study. |
| 7 | Clearly define all outcomes, exposures, predictors, potential confounders, and effect modifiers | Methods (Participants), Paragraphs 3-5; Methods (Outcomes); Methods (Statistical methods and analysis), Paragraph 3 | “An Ambulance:Escalation Index was then constructed, by dividing the total ambulance escalation area time by the total escalation area time for each department, giving a ratio describing the relative use of ambulance escalation areas for each department, regardless of the total burden of escalation area care.”  “The primary outcome was 28-day all-cause mortality among patients admitted from the ED, with the site-level Ambulance:Escalation Index as the exposure.”  “The model adjusted for patient age (natural cubic spline with four degrees of freedom), sex, hospital trauma-receiving designation… recruitment snapshot, site admission rate, and the site's total burden of escalation area care…” |
| 8 | For each variable of interest, give sources of data and details of methods of assessment (measurement) | Methods (Participants), Paragraphs 3-5; Methods (Data collection) | “There is no universally agreed definition for an escalation area. The following definition was provided to sites: ‘any area not routinely used unless the capacity of the usual ED geographical footprint is exceeded.’ Sites were asked to assign each escalation area to one of the following categories…”  “Local study teams prospectively identified all patients present in the ED during snapshots. They subsequently used electronic health records (EHR), department management systems and in-department observation to determine patients’ disposition from the ED… Data were entered using REDCap electronic data capture tools.” |
| 9 | Describe any efforts to address potential sources of bias | Methods (Statistical methods and analysis), Paragraphs 2-3 & 5; Discussion (Limitations) | “Spearman's rank correlation was used to assess whether a site's relative use of ambulance escalation areas was associated with its total burden of escalation area care… to establish that the index measures the composition of escalation care rather than its volume.”  “…this adjustment means the Ambulance:Escalation Index is independent of total escalation burden… A directed acyclic graph demonstrating the assumed causal structure is available as Supplementary Figure 1.”  “The cohort was restricted to admitted patients, if ambulance pressures alter admission thresholds… this restriction could introduce selection bias.” |
| 10 | Explain how the study size was arrived at | Methods, Paragraph 1; Methods (Statistical methods and analysis), Paragraph 5; Results, Paragraphs 1-2 | “Hospitals reporting no escalation area use were excluded.”  “The study included 134 EDs, of which 131 (97.8%) reported the use of any escalation area at any time and so were included for analysis… 18,744 patients, admitted from the Emergency Department, were included.” |
| 11 | Explain how quantitative variables were handled in the analyses | Methods (Participants), Paragraph 5; Methods (Statistical methods and analysis), Paragraphs 1 & 3 | “The mean total escalation area time for each ED was calculated in hours, as the sum of all patients’ time in any escalation area for that department, divided by the number of patients in each ED across all snapshots.”  “The characteristics of the included EDs, and patients recruited, were described as counts, or median and interquartile range (IQR).”  “…patient age (natural cubic spline with four degrees of freedom)… Hazard ratios (HRs) for the index are presented per 5 percentage-point absolute increase for interpretability.” |
| 12(a) | Describe all statistical methods, including those used to control for confounding | Methods (Statistical methods and analysis), Paragraphs 3-4 | “The association between the Ambulance:Escalation Index and 28-day all-cause mortality was estimated with a Cox proportional hazards model… The model adjusted for patient age… sex, hospital trauma-receiving designation… recruitment snapshot, site admission rate, and the site's total burden of escalation area care…”  “The baseline hazard was stratified by diagnosis group. As the exposure varies only between sites, inference was based on robust standard errors clustered at the hospital level.” |
| 12(b) | Describe any methods used to examine subgroups and interactions | Methods (Statistical methods and analysis), Paragraph 3; Results, Table 1 | “The exposure was parameterised in two parts: a binary indicator of any use of ambulance escalation areas by the site, and a continuous dose term, the Ambulance:Escalation Index.”  “The characteristics of the EDs and admitted patients, stratified by the Ambulance:Escalation Index, are presented in Table 1.” |
| 12(c) | Explain how missing data were addressed | Methods (Statistical methods and analysis), Paragraph 5 | “The data were analysed on a complete-case basis and statistical significance was set at p<0.05.” |
| 12(d) | If applicable, explain how loss to follow-up was addressed | Methods (Data collection) | “At 28-days, hospital and GP records were examined for evidence of patient death.”  All admitted patients were followed for the full 28-day period through linked hospital and GP records; there was no loss to follow-up requiring separate handling. |
| 12(e) | Describe any sensitivity analyses | Methods (Statistical methods and analysis), Paragraph 5 | “The proportional hazards assumption was assessed using scaled Schoenfeld residuals, and the functional form of continuous covariates using martingale residuals. The linearity of the dose-response association was assessed with a penalised-spline Cox model (restricted maximum likelihood smoothing) including a hospital-level random effect… The stability of the dose-response estimate was assessed by leave-one-out analysis by site.” |
| **Results** |  |  |  |
| 13(a) | Report numbers of individuals at each stage of study | Results, Paragraphs 1-2 | “The study included 134 EDs, of which 131 (97.8%) reported the use of any escalation area at any time and so were included for analysis… 37.4% (n=49) of sites reported no use of ambulance escalation areas.”  “18,744 patients, admitted from the Emergency Department, were included… 1,240 patients (6.6%) had died at 28-day follow-up.” |
| 13(b) | Give reasons for non-participation at each stage | Methods, Paragraph 1; Methods (Statistical methods and analysis), Paragraph 5; Results, Paragraph 1 | “Although part of the study reported data from 13 EDs in Scotland, due to differing national approaches to regulatory approval of recruitment via waived consent, Scotland did not collect patient level data.”  “Hospitals reporting no escalation area use were excluded.” |
| 13(c) | Consider use of a flow diagram | N/A | N/A – recruitment and participation flow for the parent cohort is reported in the primary analysis. Site-level inclusion for this secondary analysis is described in Results, Paragraph 1. |
| 14(a) | Give characteristics of study participants and information on exposures and potential confounders | Results, Paragraphs 2-3; Table 1; Figure 1 | “The characteristics of the EDs and admitted patients, stratified by the Ambulance:Escalation Index, are presented in Table 1.”  “The median age was 72 (IQR 55-83), and 9,036 (48%) were Male.”  “Figure 1. Characteristics of hospital sites, by the mean escalation time per patient, the proportion of that escalation time in ambulance…” |
| 14(b) | Indicate number of participants with missing data for each variable of interest | Methods (Statistical methods and analysis), Paragraph 5; Results, Table 1 | “The data were analysed on a complete-case basis…”  Table 1 reports complete counts for each site- and patient-level variable of interest (site designation, Ambulance:Escalation Index, mean escalation time, admission rate, age, sex and mortality). |
| 14(c) | Summarise follow-up time | Methods (Participants), Paragraph 2; Methods (Data collection) | “Demographics were recorded for all patients admitted from the ED, and these patients were subsequently screened at 28 days to determine their mortality outcome.”  All included patients contributed a fixed follow-up period of 28 days from their index ED attendance. |
| 15 | Report numbers of outcome events or summary measures over time | Results, Paragraph 2 | “1,240 patients (6.6%) had died at 28-day follow-up. Mortality was highest in those departments with highest Ambulance:Escalation Index, with 8.9% mortality for ED admissions in those departments using ambulances for >15% of their total escalation area time, compared to 6.4% for those without ambulance escalation area use.” |
| 16(a) | Give unadjusted estimates and, if applicable, confounder-adjusted estimates and their precision | Results, Paragraph 5; Table 2 | “There was no association between the binary use, or not, of ambulance escalation areas and mortality (HR 0.957, 95% CI 0.808-1.134, p=0.612). Among sites using ambulance escalation areas, increasing relative use was associated with increased mortality: each 5 percentage-point absolute increase in the Ambulance:Escalation Index was associated with a 2.1% relative increase in the hazard of death (HR 1.021, 95% CI 1.002-1.041, p=0.032).”  Table 2 presents the full output of the univariable and multivariable Cox proportional hazards model, with adjusted hazard ratios and 95% confidence intervals for every covariate. |
| 16(b) | Report category boundaries when continuous variables were categorised | Results, Table 1 | “Ambulance:Escalation Index: No Ambulance Use; >0–5%; >5–10%; >10–15%; >15%” |
| 16(c) | If relevant, consider translating estimates of relative risk into absolute risk | Results, Paragraph 2 | “…8.9% mortality for ED admissions in those departments using ambulances for >15% of their total escalation area time, compared to 6.4% for those without ambulance escalation area use.” |
| 17 | Report other analyses done | Results, Paragraphs 4-5; Supplementary Figures 2 & 3 | “For those sites using ambulance escalation, there was no significant correlation between a site's relative use of ambulance escalation areas and its mean total escalation area time (Spearman's rho = -0.149, p=0.183), supporting the index as a measure of the composition of escalation care rather than its overall burden.”  “The linearity of this relationship was confirmed with visual inspection of the penalised-spline Cox model and was robust to leave-one-out analysis (Supplementary Figures 2 & 3).” |
| **Discussion** |  |  |  |
| 18 | Summarise key results with reference to study objectives | Discussion (Statement of principal findings), Paragraph 1 | “In this secondary analysis of 131 UK EDs, the use of ambulances as escalation areas was widespread… Among sites that did use ambulance escalation area care, mortality rose steadily with the share of escalation care delivered in ambulances: each 5 percentage-point increase in the Ambulance:Escalation Index was associated with a 2.1% relative increase in the hazard of 28-day mortality among admitted patients.” |
| 19 | Discuss limitations of the study, taking into account sources of potential bias or imprecision | Discussion (Limitations) | “This is an observational between-site comparison, and because the exposure is a site-level characteristic, no within-site design is possible. Unmeasured differences between sites, particularly in staffing… Equally, other operational factors, such as estate footprint or flow management culture could confound the association… A patient was only deemed to be in an ambulance-based escalation area if their wait exceeded 15 minutes, with no equivalent threshold applied to in-hospital escalation time… it will tend to understate ambulance use at sites with frequent shorter holds.” |
| 20 | Give a cautious overall interpretation of results | Discussion (Statement of principal findings), Paragraph 2; Discussion (Strengths and weaknesses in relation to other studies); Discussion (Meaning of the study); Conclusion | “In this secondary analysis, a causal relationship has been established for neither, but the consistency and dose-dependence of the present association across 131 sites strengthens the case that the choice between ambulance and in-hospital escalation is not neutral and deserves further study.”  “Finally, this remains a hypothesis-generating secondary analysis of a study designed for a different primary question, and causal claims are not supported.”  “These results provide early evidence for policies that aim to relieve the burden of ambulance ramping. They are not, however, an endorsement of in-hospital corridor care.” |
| 21 | Discuss the generalisability (external validity) of the study results | Discussion (Statement of principal findings), Paragraph 2; Discussion (Limitations); Discussion (Meaning of the study) | “Its strengths include national scope, prospective recruitment across prespecified representative snapshots, 28-day follow-up through hospital and GP records, and an analysis designed for a site-level exposure…”  “These findings have implications across the international response to ED crowding. For example, current UK guidance requires ambulances to offload within 45 minutes, shifting escalation burden from ambulances into hospitals.”  “The cohort was restricted to admitted patients… this restriction could introduce selection bias.” |
| **Other information** |  |  |  |
| 22 | Give the source of funding and the role of the funders | Title Page (Funding) | “The study was funded by the Royal College of Emergency Medicine. Grant number RCEM24_SG_4” |

*Give information separately for cases and controls in case-control studies and, if applicable, for exposed and unexposed groups in cohort and cross-sectional studies.

**Note:** An Explanation and Elaboration article discusses each checklist item and gives methodological background and published examples of transparent reporting. The STROBE checklist is best used in conjunction with this article (freely available on the Web sites of PLoS Medicine at http://www.plosmedicine.org/, Annals of Internal Medicine at http://www.annals.org/, and Epidemiology at http://www.epidem.com/). Information on the STROBE Initiative is available at www.strobe-statement.org.
