## Supplement for "Ambulance or corridor? The association between site-level use of ambulance ‘ramping’ as Emergency Department escalation areas and 28-day mortality in admitted patients: a secondary analysis of the UNCORKED study"

Supplementary Figure 1. Directed acyclic graph demonstrating the modelling approach used to determine the association between the site-level Ambulance:Escalation Index and mortality.

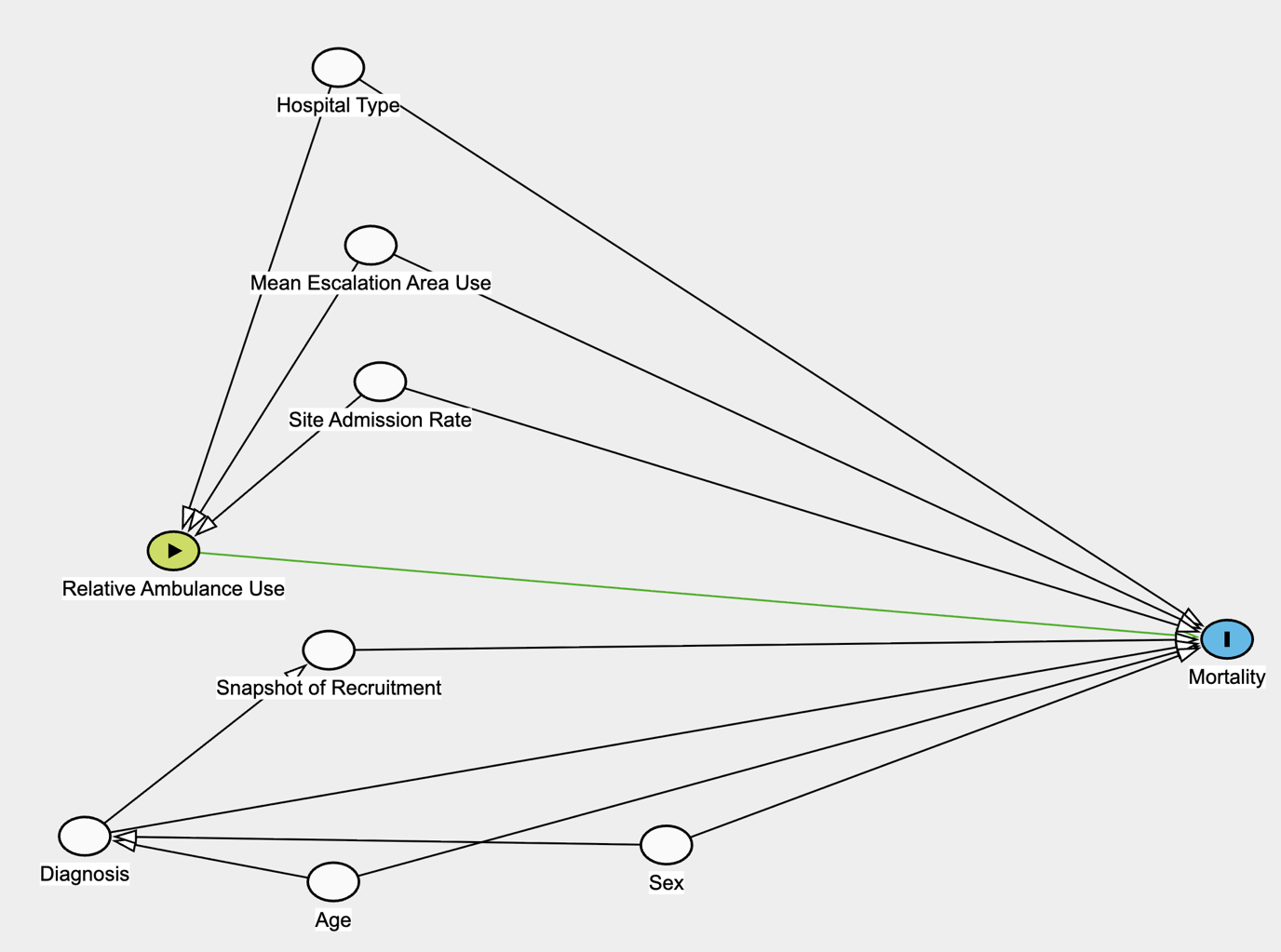

Supplementary 2. Partial effects plot for the penalised-spline Cox model assessing the linearity assumption for the association between the Ambulance:Escalation Index and mortality. Ticks indicate site-level data points.

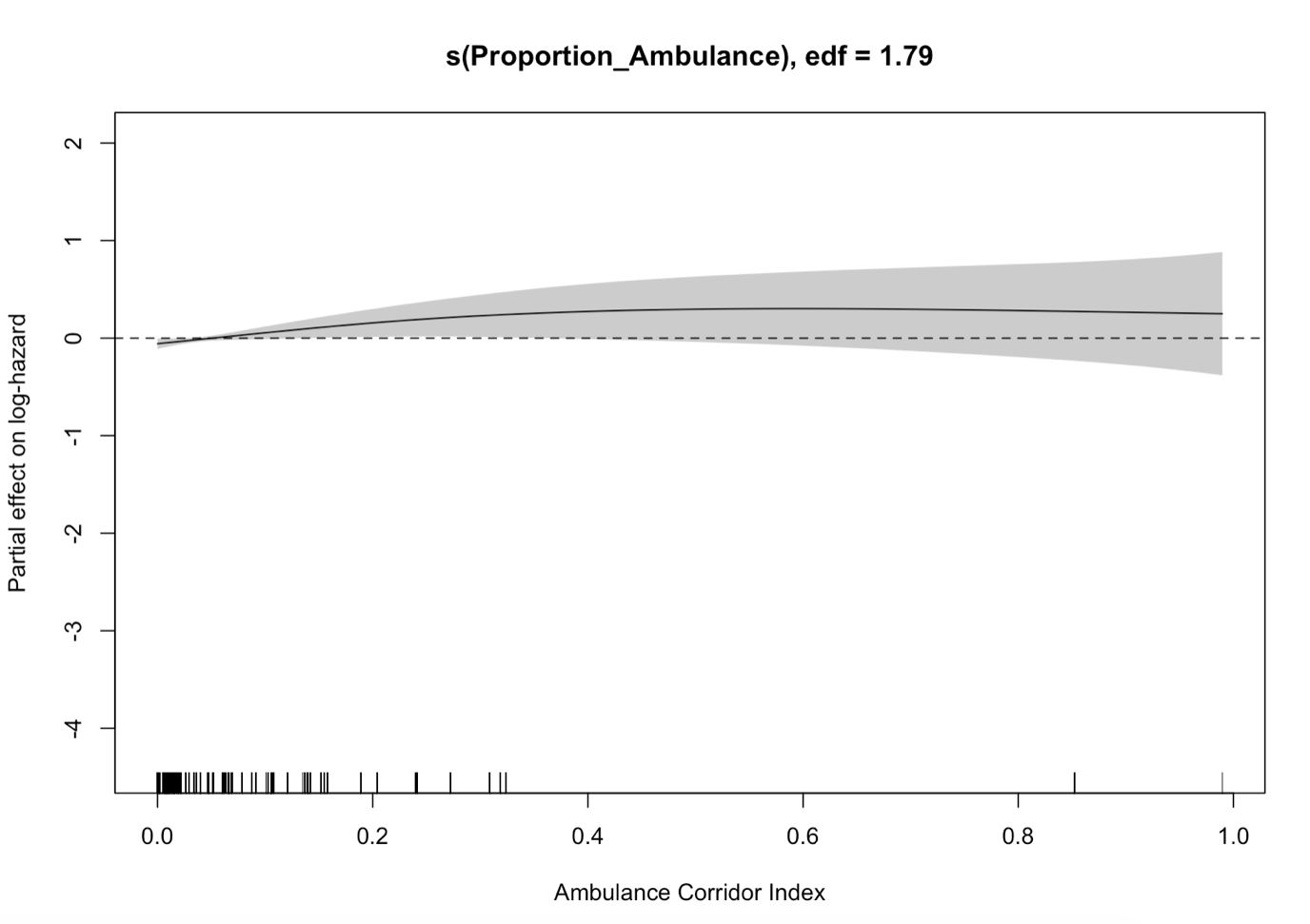

Supplementary Figure 3. Plot for leave-one-out analysis, demonstrating stability of results to single site-level influence.

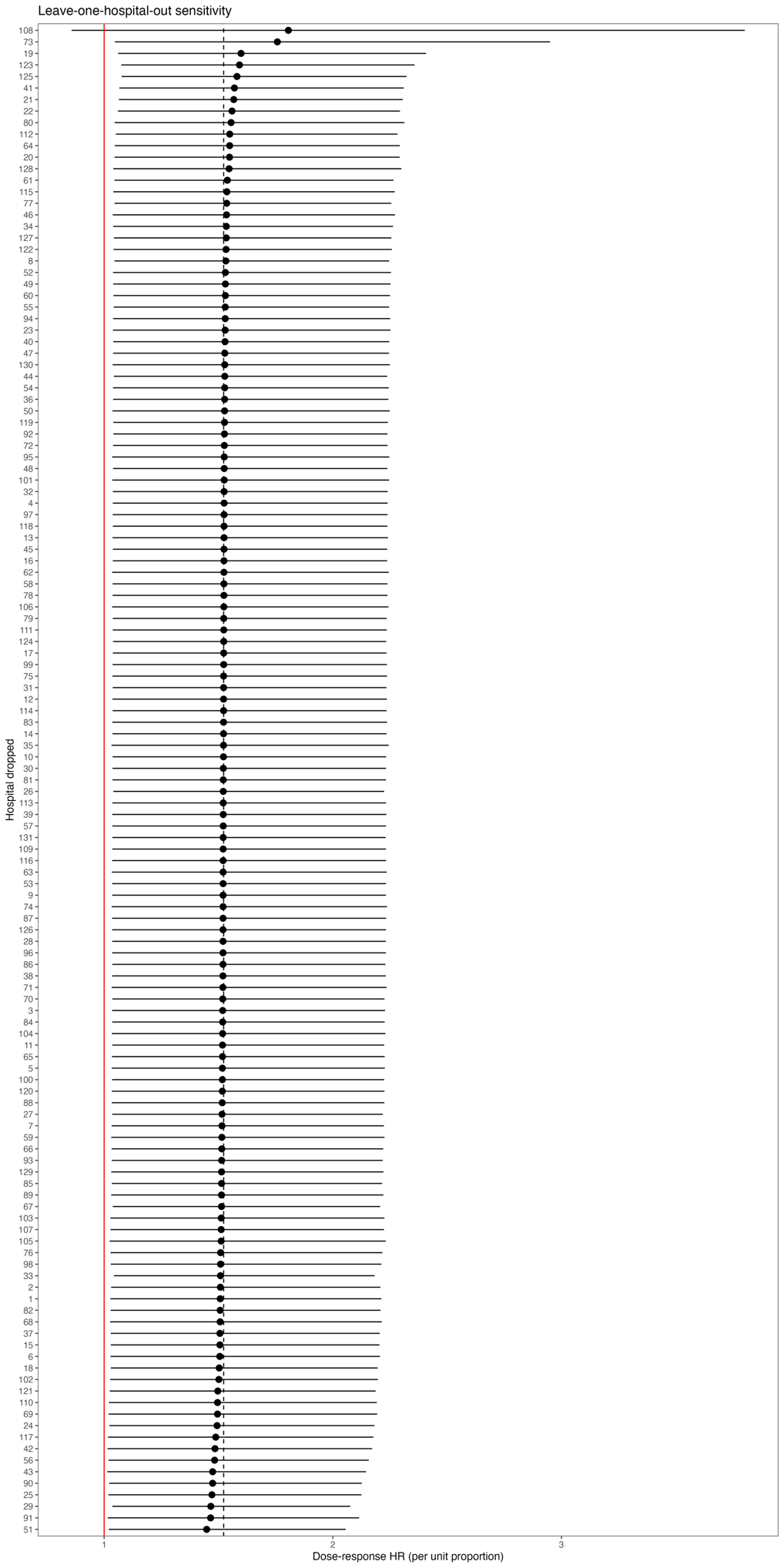
